# An empirical analysis of what people learned about COVID-19 through web search and the impacts on misinformation and attitude towards public health safety guidelines

**DOI:** 10.1101/2021.02.23.21252323

**Authors:** Ikpe Justice Akpan, Obianuju Genevieve Aguolu, Asuama Akpan

**Affiliations:** Department of Management and information Systems, Kent State University, New Philadelphia, OH 44663, USA; Infectious Disease Internal Medicine Department, Yale School of Medicine, Yale University, New Haven, CT 06510, USA; Research and Development, Ibom International Center for Research and Scholarship, Windsor, ON N8R 1A2, Canada

**Keywords:** Internet, novel coronavirus, SARS-CoV-2, COVID-19, misinformation, conspiracy theories, effective health communication strategies

## Abstract

Several people flocked to the Internet to learn about the SARS-CoV-2 and COVID-19 after the outbreak in Wuhan, China, in December 2019. As the novel coronavirus spread rapidly worldwide and was declared a global pandemic, the public rushed to Internet platforms to learn about the outbreak through Google search, online news outlets, and social media platforms. This paper evaluates the public’s web search to learn about the pandemic and the possible impacts on attitude to the public health guidelines. The results highlight four outcomes: First, a significant global population learned about the ongoing pandemic through a web search. Second, there is a direct correlation between learning SARS-CoV-2, COVID-19, and SARS-CoV and searching information on public health measures (wearing a facial mask and social distancing). Third, learning conspiracy theories or misinformation correspond with a lack of interest in gaining knowledge about public health safety guidelines. Also, the initial high interest in learning about Influenza declined as people gained information about SARS-CoV-2 and COVID-19. The results highlight the critical need to promptly sensitize the public about global health concerns using both the Internet platforms and traditional sources, adopt effective health communication strategies, and build trust.

## 1. Introduction

The use of the Internet and other Information Systems and platforms to obtain public health information or manage health-related issues has become widespread in the current digital age (Lima-Pereira, Bermúdez-Tamayo, & Jasienska, 2012; Agarwal et al., 2011). Generally, the practice has become so pervasive that the first reaction to obtaining any health information is to ‘google it’ (Lee, 2016; Chung et al., 2012). While seeking knowledge through web search is welcome, the prevalence of fake news and erroneous information on the Internet platforms raises serious concerns (Ciampaglia et al., 2018).

A novel (n) coronavirus (CoV) initially named 2019-nCoV emerged in Wuhan, China, and was formerly reported to the WHO on December 31, 2019 (Taylor, 2020; Huang et al., 2020; Wu et al., 2020). Further scientific evidence soon unveiled about the semblance of the 2019-nCoV’s genome sequence with a previous epidemic, the “severe acute respiratory syndrome” (SARS), a disease epidemic caused by SARS coronavirus (SARS-CoV), which broke out in Foshan, China, in 2002 (Shih & Sun, 2020; Chan et al., 2020; Woo et al., 2019). Some initial studies also identified similar features that relate to the “Middle East respiratory syndrome” (MERS) epidemic caused by the MERS coronavirus (MERS-CoV) as the causative agent (WHO, 2020).

The outbreak was formerly named 2019 novel coronavirus (2019-nCoV) on January 13, 2020, the same day that the first imported case occurred outside China, which took place in the Philippines and other countries (Labrague et al., 2020). The spread continued across many countries, causing the WHO to declare the outbreak a pandemic (WHO, 2020). The 2019-nCoV was later renamed SARS-CoV-2 and identified as the causative agent of the coronavirus disease 2019 (COVID-19) in February 2020 (WHO, 2020c; Atzrodt et al., 2020). The highly contagious COVID-19 continued to spread rapidly globally and caught the world unprepared. With no adequately planned health communication strategies, panic ensued, while confirmed cases of infections and deaths from COVID-19 increased astronomically worldwide (Wu, Leung, & Leung, 2020).

After sensing the potential global spread of the outbreak, the WHO launched a free online introductory training course in different languages (including English, French, Spanish, and Chinese) to sensitize people about the contagious COVID-19 (WHO, 2020). However, it is unclear how many people knew or utilized the free training lessons about COVID-19 that WHO had made available via its website (WHO, 2020). Instead, several studies suggest that the public flocked to the Internet to learn about SARS-CoV-2 and COVID-19 through a web search, Online news outlets, and social media (Allington et al., 2020; Hornik et al., 2020). The Internet’s use to learn or gain information about COVID-19 in this digital age is not surprising as Internet use has become pervasive (Akpan, Soopramanien, & Kwak, 2020). Several studies have examined social media’s influence on what people learned and the appropriate behaviors towards misinformation and conspiracy theories (Miller et al., 2020a; 2020b). Similarly, Sulyok, Ferenci, & Walker (2020) examine the impact of web search on the confirmed cases of COVID-19 in Europe (Sulyok, Ferenci, & Walker, 2020). This paper becomes the first to undertake an empirical investigation on using a “web search” to learn about SARS-CoV-2 and COVID-19 and learners’ attitude to public health guidelines.

Miller et al. (2020a; 2020b) identify political leaders’ failure in sensitizing the public as another important reason people considered the Internet as an information source alternative to learning about the pandemic. Misinformation had started flooding the web right from the initial stage of the emergence of the novel coronavirus, mainly from user-created content on social media (Bautista, Zhang, & Gwizdka, 2021). Thus, as people turned to the web searching for information, there was limited, non-technical information about the coronavirus. People rather got exposed to either learning wrong information about SARS-CoV-2 and COVID-19 or embrace “fake news,” “misinformation,” and conspiracy theories, with grave consequences (Allington et al., 2020). Some of the unfounded misinformation include misconstruing COVID-19 as “common cold” (Rovetta, & Bhagavathula, 2020), while the conspiracy theories propagated online include treating COVID-19 as “5G” and “bioweapon” (Garrett, 2020; Ahmed et al., 2020; Miller et al., 2020a; Miller et al., 2020b).

With no cure for the COVID-19 yet, the public health experts offer some safety guidelines and protective health behaviors, including social distancing and wearing a facial mask to limit the spread; isolation and quarantine for those with imminent risks (CDC, 2020; Hornik et al., 2020). People who learn or believe the misinformation and conspiracy theories tend to ignore public health warnings, a situation which experts consider as the reason the USA records one of the highest infection rate and fatality globally (19.97 million confirmed cases and 345,737 deaths, respectively) as of December 31, 2020 (CDC, 2020).

This paper examines what people learned about SARS-CoV-2 and COVID-19 pandemic through web search (as indicated by the Google search trends data), the possible impacts on behavioral responses to the public health guidelines, and potential consequences on the spread of COVID-19. The rest of the paper is structured as follows: the next section (two) states the research objectives, presents a theoretical background for this study. Section three discusses an overview of the SARS-CoV-2 outbreak and COVID-19 pandemic, while Section four presents the materials and method, including data collection. Section five analyzes and discusses the results. Finally, Section six highlights the paper’s contributions, policy implications of the study, and recommendations for future studies.

## 2. Research Questions and Theoretical background

### 2.1 Research Objectives

This study seeks to answer the following research questions:

a. What did people learn about SARS-CoV-2 and COVID-19 through “web search”? Here, the terms, keywords, or phrases that people used to search the web approximates what the public learned or knowledge acquired about the novel coronavirus.
b. Did what people learned impact behavior towards the public health safety measures (wearing facial mask, social distancing, washing hands, and more?
c. What are the public health implications of (a) and (b) above?

### 2.2 The Connectivism Learning Theory

This section examines the connectivism learning theory, which explains using digital platforms to enable learning (Siemens, 2005; Downes, 2008; Goldie, 2016). This study employs the approach to explore how people learned about SARS-CoV-2 and COVID-19 through ’web search’ and the potential behavioral implications towards public health guidelines, which scientists and medical experts recommend as ways to check the spread of COVID-19. E.g., we investigate if learning through ’web search’ helped people acquire accurate knowledge or misinformation and conspiracy theories about the COVID-19 pandemic and its implications. Also, recent studies show that many people are yet to understand the science and the concept of the novel coronavirus (SARS-CoV-2) and the disease, COVID-19, which increases the danger of embracing misinformation (Lee et al., 2020; Miller et al., 2020b). Several web platforms, including social media, Online news, and other Internet channels, contribute significantly to misinformation and conspiracy theories (Mian & Khan, 2020; Miller et al., 2020a).

As proposed by George Siemens (Siemens, 2005), the connective learning theory analyzes the use of digital devices, computer networks, and electronic platforms to learn. The view is considered a pedagogical strategy for the digital age, emphasizing knowledge sharing across an interconnected Web and Internet network (Goldie 2016; Siemens, 2005). The approach focuses on knowledge acquisition utilizing information technology platforms and learning from multiple sources, developing skills, and disseminating information (Downes, 2008). The platforms incorporate information on social media, Internet websites or blogs, and search engines that users can employ to learn and exchange knowledge, skills, and expertise (Dunaway, 2011; Clarà, & Barberà, 2013).

One of the theory’s implications is that learning can occur outside the traditional classroom to use networked systems that enhance connections, interactions, and collaborations among the learners (Corbett and Spinello 2020). However, some learning theory experts criticize the connectivism theory for not offering any improvement to the actual learning method other than utilizing the Web 2.0 and related platforms (Downes 2008). Hence it cannot be deemed a substantive learning theory. Instead, it provides a bridge to other pedagogical methods: behaviorism, cognitivism, and constructivism. The core of the Siemens and Downes connectivism idea aims to move away from the traditional classroom learning techniques to a new theory of learning that embraces technology as the learning tool, which can inspire the new generation of learners and educators (Dunaway, 2011). Thus, the theory draws its strength from Web-based activities (Corbett, & Spinello 2020).

The key benefit of the method is its intuitiveness and the ability to captivate learners due to the ubiquitous use of the Internet in today’s world. The following principles contribute to the popularity of connectivism as a learning theory (Siemens 2005).

i. Learning and knowledge rest in diversity of opinions, as experienced today.
ii. Learning is a process of connecting specialized nodes or information sources.
iii. Learning may reside in non-human appliances.
iv. The capacity to know more is more critical than what is currently known.
v. Nurturing and maintaining connections help to facilitate continual learning.
vi. Ability to see connections between fields, ideas, and concepts is a core skill.
vii. Currency (accurate, up-to-date knowledge) is the intent of all connectivism learning activities.
viii. Decision-making is itself a learning process. Choosing what to learn and the meaning of incoming information is seen through a shifting reality.

The connectivism learning theory, as explained above, closely mirrors the use of Google search trends and other Internet platforms to learn about the outbreak of SARS-CoV-2 and COVID-19, especially where the masses did not get adequate, timely information about the novel coronavirus from the public agencies.

The connectivism learning theory, which is well suited to personal study and self-regulated learning (Schunk, 2005; Akpan, 2014), in this case how individual members of the public learned about SARS-CoV-2 and COVID-19 using web search in the first six (6) months of the COVID-19 pandemic. An

## 3. Overview of the SARS-CoV-2 Outbreak and COVID-19 Pandemic

### 3.1 The Global Impacts of SARS-CoV-2 Outbreak

The COVID-19 pandemic has inflicted severe problems ranging from health crises to psychological, social, and business and economic consequences the world over (Miller et al., 2020b; Akpan, Soopramanien, & Kwak, 2020; Donthu & Gustafsson, 2020). Meanwhile, there is currently no specific cure for COVID-19. However, there has been significant progress and technological advances leading to substantial breakthroughs in vaccine discovery and development through the pioneer efforts by Pfizer, Madonna, and others from China and other countries (Tuite et al., 2021). Administering the COVID-19 vaccines are ongoing worldwide, while several other vaccine discoveries and developments are in progress (Wimalawansa, 2020). In the meantime, ongoing prevention, monitoring, and public health awareness are essential to mitigate the public health and economic burden. The most important prevention strategy is to understand the disease and how it spreads.

### 3.2 Etiology

SARS-CoV-2 is a member of the large coronavirus family. Coronaviruses are enveloped, positive-sense, single-stranded RNA viruses. There are four main sub-groupings of coronaviruses – alpha, beta, gamma, and delta, which causes illnesses in humans and animals (cats, cattle, camels, and bats). In contrast, human coronaviruses cause most cases of common colds (Wimalawansa, 2020). The infection is usually seasonal and commonly occurs during the winter or spring seasons. The typical human coronavirus strains are from the alpha and beta sub-groups (CDC, 2020a). Animal coronaviruses may infect humans and have previously resulted in outbreaks. The first known coronavirus outbreak was the severe acute respiratory syndrome (SARS) in 2002-2003 caused by SARS-CoV. In 2012, there was an outbreak of another coronavirus disease -Middle East Respiratory Syndrome (MERS). SARS-CoV, MERS-CoV, and SARS-CoV-2 are all beta coronaviruses. All three strains originated from bats (CDC, 2020a).

### 3.3 Transmission

The coronavirus transmission is primarily through respiratory droplets released from infected persons during cough, sneeze, or speech. One can also contact the virus via contact with contaminated surfaces. The virus can remain infectious in the air for 3 hours and on inanimate surfaces for 2 to 3 days up to 9 days or longer. This has implications for nosocomial spreads and super-spreading events (van Doremalen et al., 2020). The virus has also been isolated from blood, urine, and stool specimens. It is important to note that asymptomatic infected people may not be aware that they are infected because they do not have the symptoms or may not recognize the symptoms. Infected individuals can be contagious for up to 4 weeks and can unknowingly be spreading the infection. (van Doremalen et al., 2020).

### 3.4 Clinical Presentation and Diagnosis

Symptoms usually appear 2 to 14 days after exposure. Most confirmed cases of SARS-CoV-2 infection are asymptomatic, and they recover without treatment. Common symptoms include fever, cough, shortness of breath, chills, myalgia, headache, sore throat, anosmia, and dysgeusia. Severe cases present with dyspnea, tachypnea, hypoxia (blood oxygen saturation </=93%), the arterial partial pressure of oxygen to fraction of inspired oxygen [PaO2/FiO2] less than 300, lung infiltration (Ai et al., 2020). Some patients present with gastrointestinal symptoms such as vomiting, diarrhea, abdominal pain, and cardiovascular features such as arrhythmia, shock, and acute cardiac injury (CDC, 2020b). There have been reports of asymptomatic carriers presenting with symptoms such as loss of smell and taste. In children, the majority present with mild (fever, cough, fatigue, congestion), moderate (pneumonia) symptoms (CDC, 2020b). Some may be asymptomatic. Children < 5 years old may present with respiratory organ failure.

Chest CT scan shows a distinct appearance of ground-glass lung opacity, often bilateral, in patients who develop pneumonia (Ai et al., 2020). Other radiographic features such as “crazy-paving sign, multifocal organizing pneumonia, and architectural distortion in a peripheral distribution” may appear with disease progression. Diagnostic testing is performed from respiratory (nose, throat, saliva) and serum samples, using a real-time Reverse Transcriptase Polymerase Chain Reaction (RT-PCR) panel or antibody test. The viral RNA has also been detected in stool and blood (Woo et al., 2020).

### 3.5 Complications

Some hospitalized patients develop thromboembolism, especially deep venous thrombosis, and pulmonary embolism. Other complications include Microvascular thrombosis of the toes, clotting of catheters, myocardial injury with ST-segment elevation, and large vessel strokes. This complication may be associated with the release of high levels of inflammatory cytokines and activation of the coagulation pathway caused by hypoxia and systemic inflammation secondary to COVID-19 (NIH, 2020).

### 3.6 Prevention and Control

People must be well-informed. Infected persons must practice respiratory etiquette to avoid infecting others, including covering coughs and sneezes with a tissue and discarding it properly, coughing into the inside of the elbow, and covering the nose and mouth properly with a surgical face mask. Best practices include proper handwashing with soap and water for at least 20 seconds or at least 60% alcohol-based hand rub. Clean touched surfaces with disinfectants frequently. Avoid touching the eyes, nose, and mouth with unwashed hands. Avoid close contact with people who are ill (Soo et. al., 2020). CDC recommends that infected and exposed individuals must isolate or quarantine themselves, respectively, for at least 14 days. The CDC also recommends social distancing (avoid mass gatherings or large community events, shaking hands, or giving “high fives”) (Wimalawansa, 2020). In healthcare settings, standard contact, and airborne precautions, as well as eye protection, should be used to mitigate the spread of SARS-CoV-2 (Soo et al., 2020). There is no specific cure for COVID-19. Management is mainly supportive care and treatment of secondary infections. Severely ill patients may need advanced organ support.

## 4. Materials and Method

### 4.1 Google Trends of Search Terms about SARS-CoV-2 and COVID-19

This paper evaluates the use of web search to learn about SARS-CoV-2 and COVID-19. The study utilizes the Google trends data (www.google.com/trends), an evolving resource generated through live searches of terms, keywords, and phrases. The ’Google trends’ is gradually becoming a reliable tool for digital epidemiology (Cervellin et al., 2017) and other healthcare studies (Nuti et al., 2014; Sulyok et al., 2020). Recent studies demonstrate the validity and relevance of Google trends data in healthcare research, including investigating the impacts of behavioral use of Online searches on the ongoing COVID-19 infections, which are available elsewhere (Cervellin et al., 2017; Rovetta & Bhagavathula, 2020). Similarly, (Akpan & Akpan, 2021) utilizes the Google trends data to examine the popularity of concepts and measure temporal trends and predict the future growth and popularity of information systems research disciplines.

This study identifies and analyzed a total of twenty-five (25) keywords or phrases representing used by the public to learn about COVID-19. The search terms either directly explain SARS-CoV-2 and COVID-19 or related previous coronavirus epidemics (e.g., SARS-CoV, SARS, MERS-CoV, MERS). Other terminologies misinformation or conspiracy theories (e.g., ’China Virus,’ Common Cold, or Bioweapon). The third category of search terms related to the public health safety measures that experts recommend as guidelines to limiting the human-to-human transmission of COVID-19 (social distancing, wearing a facial mask, washing hands, and more, [Hornik et al., 2020]). We arrived at the ’web search’ terms used to learn about the ongoing pandemic by reviewing the literature and the overview of the concept and science of COVID-19 (see Section 3). Table 1 presents the complete list.

**Table 1.** The learning terms about COVID-19 pandemic, misinformation and conspiracy theories, and the public health safety guidelines.

| **Web Search/Learning Terms Used to Learn about SARS-CoV-2 and COVID-19** |  |
| --- | --- |
| **COVID-19 & Related Epidemics** | 2019-nCoV (Wu, Leung, & Leung, 2020); nCoV (Wu, Leung, & Leung, 2020; WHO, 2020c); SARS-CoV-2 (van Doremalen, 2020); COVID-19 (WHO, 2020b; 2020c; Masters-Waage, Jha, & Reb, 2020; Ahmed et al., 2020); Pandemic (WHO, 2020; Atzrodt et al., 2020); MERS-CoV (Jo et al., 2019); MERS (Mahase, 2020; Jo et al., 2019); SARS-CoV (van Doremalen et al., 2020); SARS (Mahase, 2020); Virus (WHO, 2020b); Coronavirus (Huang et al., 2020; WHO, 2020a; Mahase, 2020); Influenza (Li, et al. 2020) |
| **Misinformation/Conspiracy Theories** | Virus Hoax (Miller et al., 2020a; 2020b); Injecting/Ingesting Bleach (Chary et al., 2020); 5G technology enhancing the spread of the virus (Ahmed et al., 2020; Fleming, 2020); COVID-19 Hoax (Garrett, 2020; Miller et al., 2020a; 2020b); Common Cold: Cold2020 (Rovetta, & Bhagavathula, 2020); China Virus (Masters-Waage, Jha, & Reb, 2020); Deliberately Released (Miller et al., 2020a); Bioweapons created by China (Miller, et al., 2020a; Ahmed et al., 2020) |
| **Public Health Measures** | Social Distancing (Matias, Dominski, & Marks, 2020); Washing of Hands or Wash Hands (Matias, Dominski, & Marks, 2020); Wearing a "Facial Mask" (Leung, Lam, & Cheng, 2020); Isolation (Matias, Dominski, & Marks, 2020); Quarantine (Matias, Dominski, & Marks, 2020) |

The Google trends (www.google.com/trends) data can be mined based on search keywords, trending topics, or search terms on a country’s basis or as the worldwide aggregate search index. The Google trends hold the trending “web search” data like search volume. The recorded search index in each period indicates the popularity of the terms. The maximum daily, weekly, or monthly value is 100, representing a 100% popularity or interest in the subject for the specified period. In contrast, the search index of 50 implies half popularity, while zero (0) means no interest or not enough data to determine the term’s popularity (Google.com/trends). This study utilizes the worldwide aggregate index bearing in mind the global impact of the covid-19.

### 4.2 Research Hypotheses

Internet platforms play a significant role in health communication during the ongoing COVID-19 pandemic. Several studies attribute the increase in misinformation and conspiracy theories about COVID-19 in different countries to the use of web platforms to learn about SARS-CoV-2 and COVID-19, including web search, social media use, and online news media (Kim, Ahn, Atkinson, & Kahlor, 2020; Rovetta & Bhagavathula, 2020). However, most of the studies are anecdotal with no empirical evidence. This study employs an empirical analysis technique.

Studies examining learning by “web search” emphasizes the significance of the search term or phrases to what the users intend to learn. Further, a trending term on the web indicates what information people are interested in learning (Roscoe et al., 2016; Rovetta & Bhagavathula, 2020; Hölscher & Strube, 2000). Based on the above premise, we formulate three (3) research hypotheses to address the study’s objectives listed in Section 2.1.

#### Hypothesis #1

The purpose of the first hypothesis is to determine what people learn about the COVID-19 pandemic through “web search.” The null and alternative hypotheses are defined thus:

- H1_0_: People did not gain useful knowledge about the ongoing COVID-19 global health pandemic.
- H1_1_: People gained useful knowledge about the ongoing COVID-19 global health pandemic.

#### Hypothesis #2

Based on the literature, using a “web search” to learn about a subject of interest can influence the learner’s decision-making and actions (Roscoe et al., 2016). On this premise, the study examines any association between what people learned about COVID-19 and people’s behavior towards the public health guidelines. In addressing the above discourse, we develop two separate hypotheses relating to “web search” to learn about COVID-19 (concept, science, and structure of SARS-CoV-2 and COVID-19), misinformation and conspiracy theories, and the behavioral response to the public health measures. The null and alternative hypotheses are as follows:

#### Hypothesis #2

- H2_0_: There is no association between what people learn about COVID-19 through “web search” and behavior towards public health measures.
- H2_1_: There is an association between what people learn about COVID-19 through “web search” and behavior towards public health measures.

#### Hypothesis #3

The widely held assertion that misinformation and conspiracy theories about the COVID-19 pandemic have had a significant impact on people’s behavior towards the public health measures.

- H3A_0_: There is no association between misinformation learned about COVID-19 and peoples’ behavior towards public health measures.
- H3A_1_: There is an association between misinformation learned about COVID-19 and peoples’ behavior towards public health measures.

Similarly, we defined the null and alternative hypotheses for learning about conspiracy theories thus:

- H3B_0_: There is no association between conspiracy theories learned about COVID-19 and peoples’ behavior towards public health measures.
- H3B_1_: There is an association between conspiracy theories learned about COVID-19 and peoples’ behavior towards public health measures.

### 4.3 Data Analysis

The statistical analysis involves evaluating relationships among the listed variables using the statistical trends through graphical display and correlation analysis. We use the statistical analysis package called JMP, one of the SAS statistical software packages (Freund & Littell, 1986), and Microsoft Excel to undertake the statistical analyses, including creating the charts, graphs, and computing the correlation matrix. The evaluation helps establish the direction of the relationships among the variables, which can be direct or inverse. A direct association occurs when two variables (dependent and independent variables) move in the same direction. On the other hand, an inverse relationship indicates that when one variable moves upward, the second variable moved in the opposite direction.

## 5. Analysis of Results and Discussion

The data used for this study come from Google trends worldwide data and covers the period from the initial outbreak of the SARS-CoV-2 in December 2019, up to June 2020, when the COVID-19 pandemic became more widely known (Huang et al. 2019). The reason for capturing the first six (6) months of the pandemic (January 1, 2020, to June 30, 2020) is capturing what people learned during the initial months, study the extent to which the self-regulated learning through “web search” influenced individual’s actions towards the public health safety measures. In recent, the lingering effect of the pandemic makes people increasingly aware of COVID-19. Also, the viral outbreak has caused severe social problems for friends and families (Lebow, 2020; Akpan & Ezeume, 2020), loss of business and economic decline (Akpan, Udoh, & Adebisi, 2020), in addition to health crises (Matias, Dominski, & Marks, 2020).

### 5.1 What Did People Learn about COVID-19 through Web Search?

The search terms and keywords that people utilized to learn about the COVID-19 pandemic through web searches point to what they learned (Hölscher & Strube, 2000). Table 1 (previous Section) identified twelve (12) learning terms as related to the ongoing SARS-CoV-2 and COVID-19. However, nine (9) of the keywords (SARS and SARS-CoV, MERS and MERS-CoV, Influenza, Virus, Pandemic, and Coronavirus) existed before the SARS-CoV-2 and COVID-19 era. The remaining four are names coined at different times by WHO for the ongoing global health crisis (2019-nCoV & nCoV for short, SARS-CoV-2, and COVID-19). Using information about previously known epidemics to learn about new outbreaks is not unique in healthcare analysis. Previous studies (e.g., Van Doremalen et al., 2020) show that people often utilize historical time-trends healthcare analysis of previous diseases to investigate new or recent disease epidemics (Van Doremalen et al., 2020). Further, WHO initially associated the outbreak of 2019-nCoV to previous epidemics, namely, SARS-CoV and MERS-CoV, the two causative agents of the last two coronavirus diseases, SARS, MERS, respectively (as explained in the previous Section). Figure 1 shows a smooth monthly moving average of the “web search” trends of all the twelve (12) keywords used by the public to learn about the COVID-19 pandemic. The early days in January 2020 show terms relating to previous epidemics, particularly MERS, SARS, Influenza, and MERS-CoV and SARS-CoV used to learn about the pandemic. This occurred prior to naming the novel coronavirus, 2019-nCoV on January 13, 2020, as explained in the previous Section.

**Figure 1.**
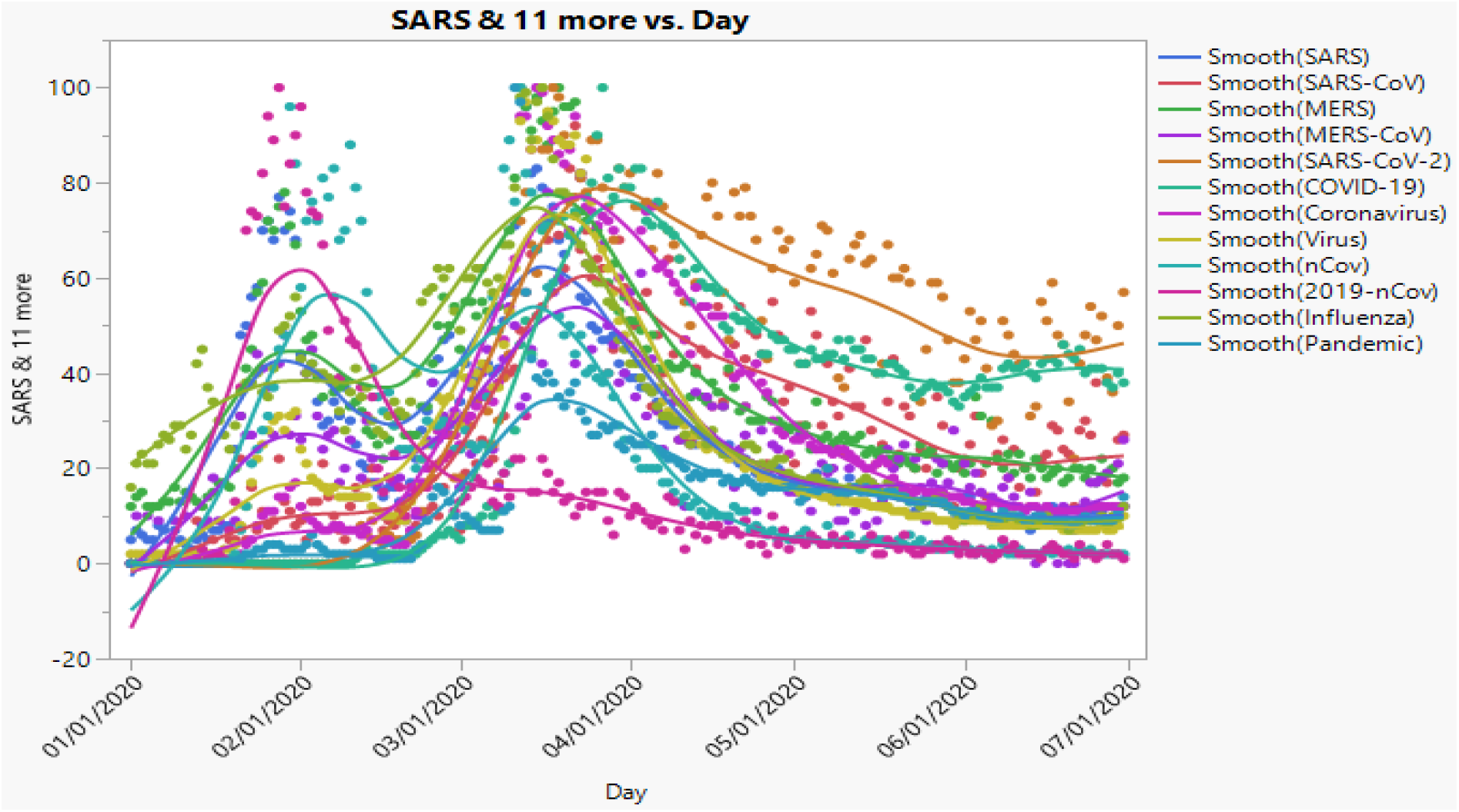
Monthly moving average of worldwide daily web search index of terms used to learn about SARS-CoV-2 and COVID-19 through (January 1, 2020 to June 30, 2020)

Overall, the three most used search keywords during the period were Influenza, MERS, and SARS-CoV-2, based on the average daily search index of 32.7, 37.2, and 39.8, respectively, out of a maximum index of 100 (according to the Google trends). Also, COVID-19 was the next most used word (4th) with an index of 32.4 (Figure 2). Despite the late start in using SARS-CoV-2 to learn about the pandemic after renaming the 2019-nCoV, it still came up as the most popular learning term. The learning term used in the early days of the outbreak was 2019-nCoV or nCoV (for short), and the least used words with an average daily search index of 15.6 and 21.9, respectively. The low popularity of these two keywords was somewhat surprising, being the coronavirus’s first official name. However, the two search terms refer to the same thing, but people tend to prefer nCoV (for short) compared to 2019-nCoV. Thus, combining both words’ usage would make it the second most popular learning search keywords (37.5) after SARS-CoV-2.

**Figure 2.**
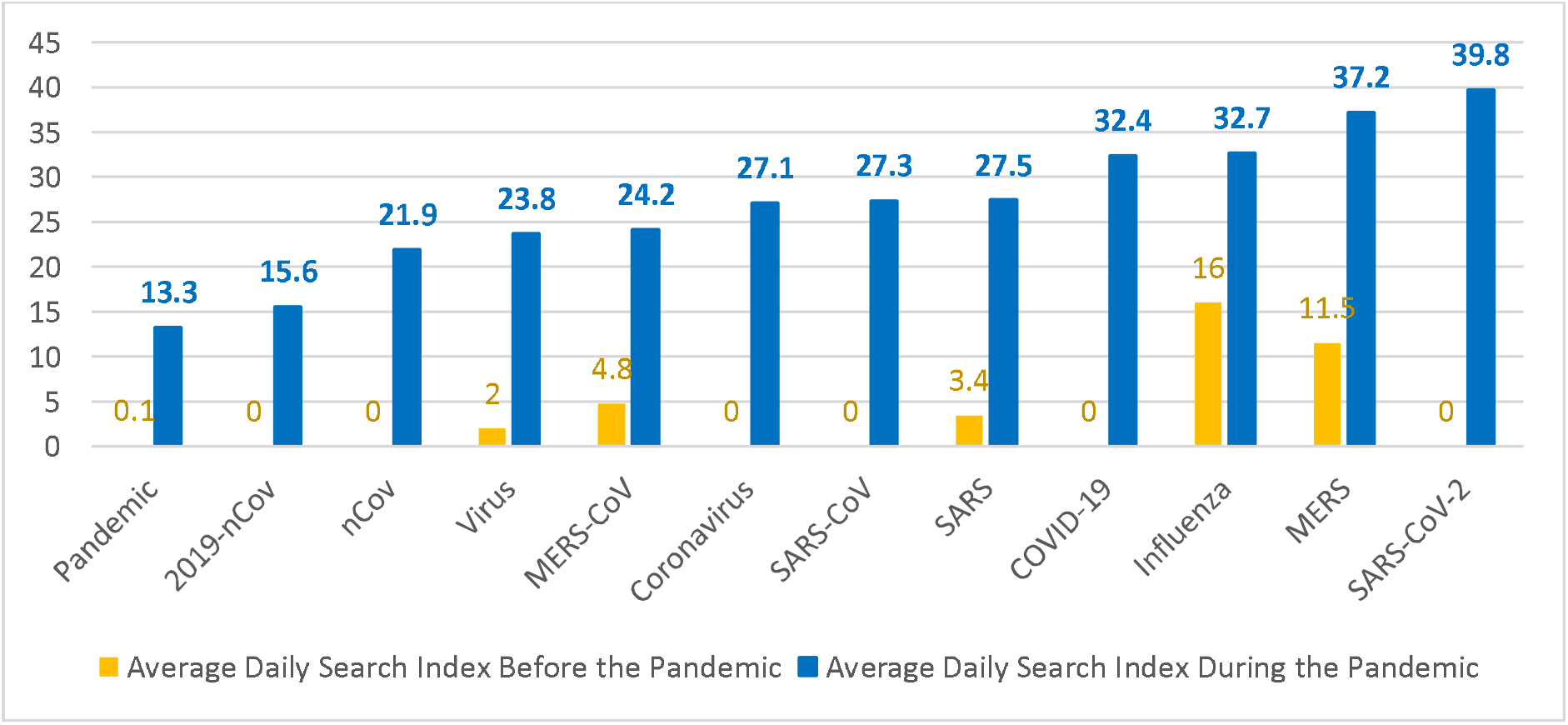
Worldwide daily average usage of the learning terms before the announcement of the outbreak on 31 December 2019 and during the COVID-19 pandemic till 30 June 2020

Some of the search keywords pre-existed before the outbreak of SARS-CoV-2 and COVID-19. For example, the learning terms (SARS, SARS-CoV, MERS and MERS-CoV, Influenza, Virus, Pandemic, and Coronavirus) refer to previous epidemics. It is plausible to argue that using these pre-existed keywords relates to ‘web search’ for purposes other than learning about COVID-19. Based on this assumption, we conduct a dependent paired t-test sample to determine if the mean search index during the period were indeed to learn about COVID-19. We compare the mean search index before and after the outbreak for each term (Figure 2). The null hypothesis for the search index of each learning term (SARS and SARS-CoV, MERS and MERS-CoV, Influenza, Virus, Pandemic, and Coronavirus) before and after the outbreak of the ongoing COVID-19 is less than or equal to zero (H_0_: µ_d_ ≤ 0) and the alternative hypothesis as (H_1_: µ_d_ > 0). The results show high variances and significant differences between the mean daily usage (> 60% in all cases) for each learning term. With clear evidence in all cases to conclude that µ_d_ > 0, we reject the null hypotheses. The daily usage of the pre-existing learning terms was indeed to learn about SARS-Cov-2 outbreak and COVID-19.

Similarly, the search term “common cold,” identified as one of the misconceptions about COVID-19 (Miller et al., 2020a; Table 1), also existed before the COVID-19 outbreak. Like other search terms discussed above, we investigate the mean search index before (cold2019) and after the outbreak (cold2020) to ascertain if the ’web search’ was to learn about the ongoing pandemic. Figure 3 shows the search index trend during the COVID-19 (Jan 1, 2020, to Jun 30, 2020). On the other hand, the search index before the outbreak (Jan 1, 2019, to Jun 30, 2019) covering the same period in the past year.

**Figure 3.**
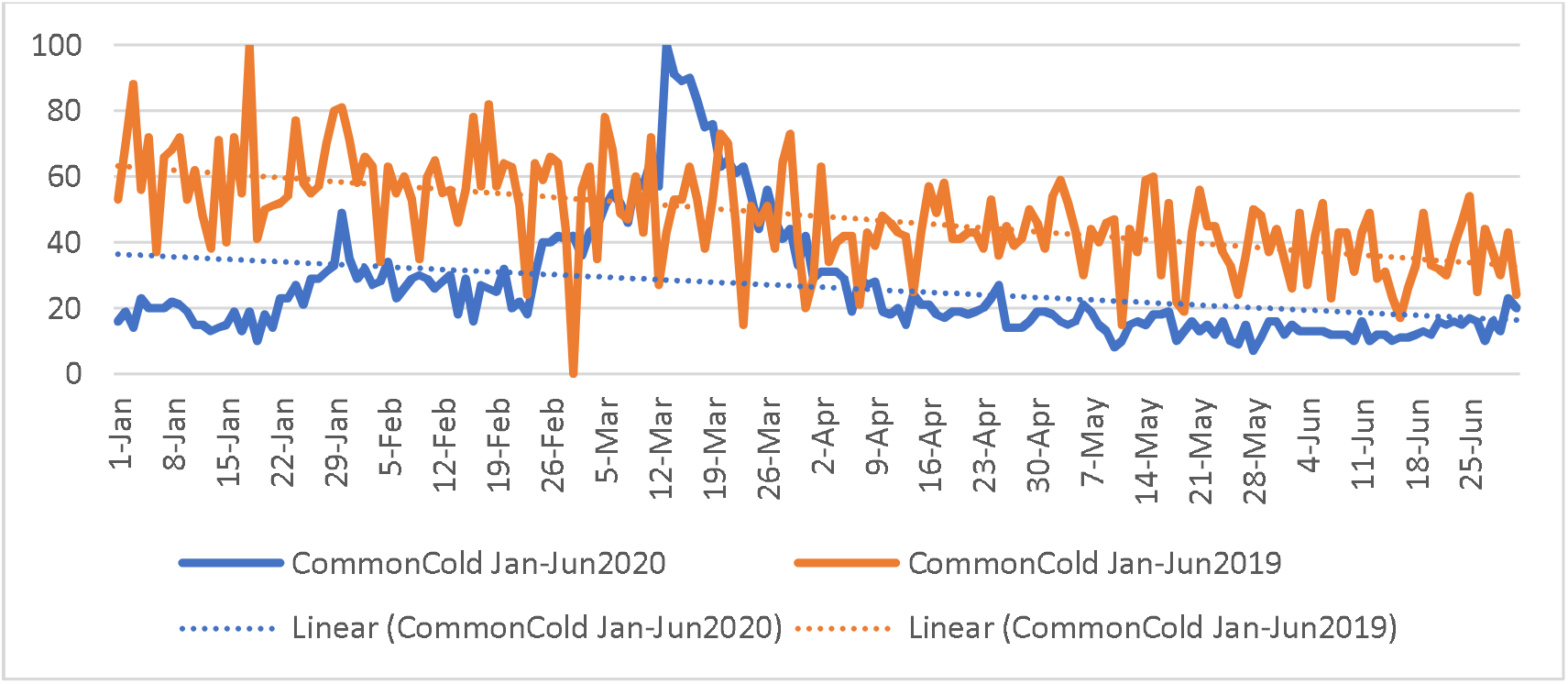
Daily web search index for the term “common cold” (cold2019, cold2020) covering the pre- and during the COVID-19 (Jan-Jun2019 v Jan-Jun2020).

The result shows a significant difference between the mean daily search index for the “common cold” learning term before (cold2019: 46.0) and during the pandemics (cold2020: 26.4) daily. Therefore, we reject the null hypothesis of the equality of mean (H_0_: µ_d_ ≤ 0) and accept the alternative view that the mean search index for the term before COVID-19 and during the pandemic (H_1_: µ_d_ > 0). is statistically different. We conclude that the difference in the average daily search index was related to the issues about COVID-19. However, unlike the other variables, the search index for “Common Cold” declined during the COVID-19 pandemic. This is an unexpected result, which tends to corroborate several studies reporting the unintended positive consequences of COVID-19 across the world, concluding that “common cold” or “flu” cases tend to witness a decline during the COVID-19 (Soo et al., 2020). Thus, exercising the public health safety guidelines of wearing facial masks and social distancing has possibly helped reduce the spread of “common cold.”

#### 5.1.1 What Search Term Contributed to Learning about SARS-CoV-2 and COVID-19?

Figure 2 presents the average search index for each search term used to learn about SARS-CoV-2 and Covid-19. In this section, we evaluate the terms that contribute to learning using the coefficient of determination. The result (Figure 4) shows that, out of the twelve (12) learning keywords, only seven (7) somewhat contributed to the learning through ‘web search.’ The coefficient of determination (R^2^) of the variables are (SARS-CoV-2: 0.37; COVID-19: 0.36; Influenza: 0.27; 2019-nCoV: 0.24; nCoV: 0.18; SARS: 0.12; SARS-CoV: 0.11). The terms that did not contribute meaningfully to learning about the COVID-19 pandemic were excluded from further analyses (MERS, MERS-CoV, Pandemic, Virus, Coronavirus, all with R^2^ < 0.1). Also, nCoV and 2019-nCoV are the same terms. Hence 2019-nCoV, which contributes more to learning, was selected for further analysis (Figure 4).

**Figure 4:**
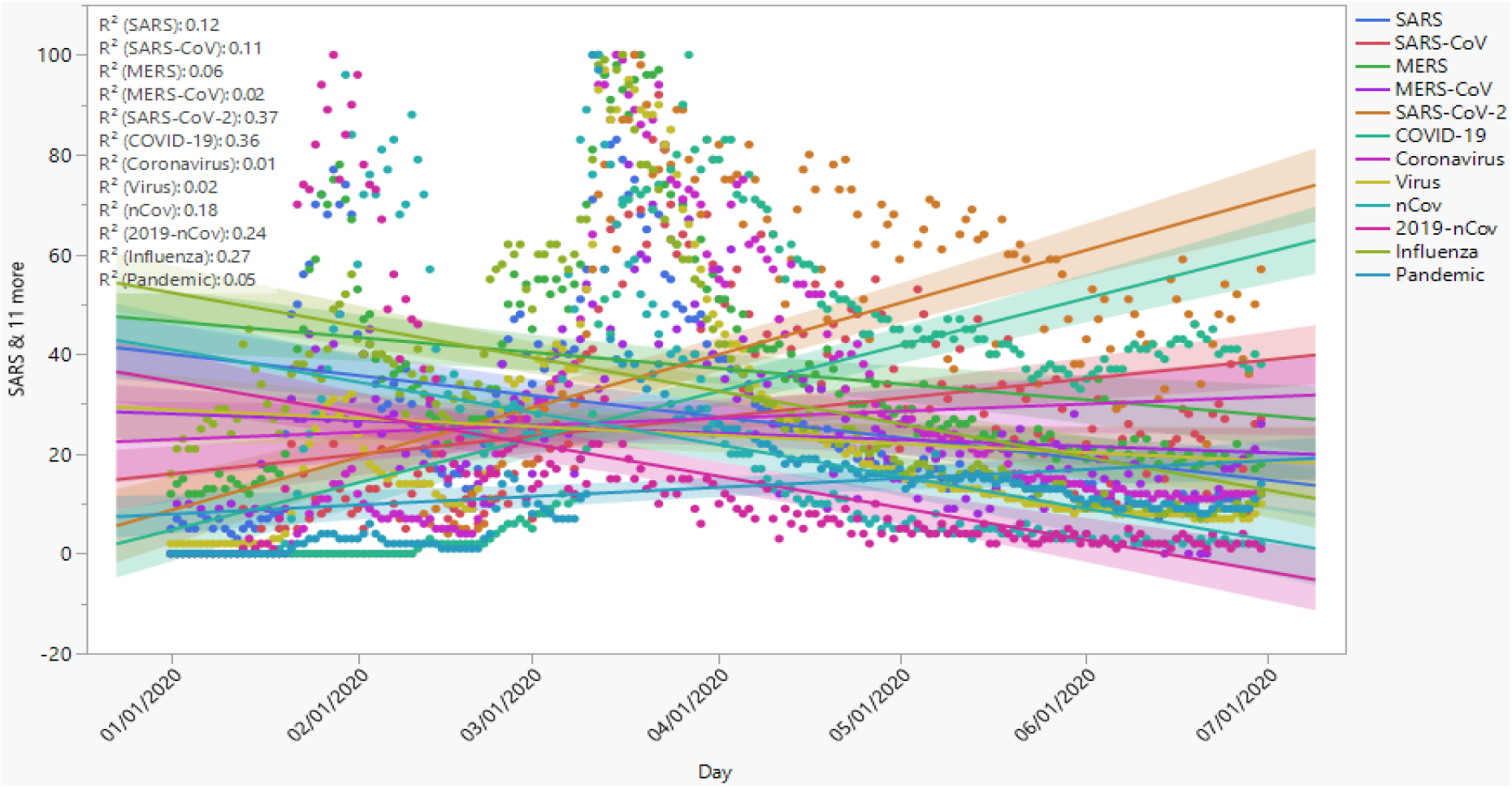
The contribution of search terms to learning about SARS-CoV-2 and COVID-19.

The surprising results are the insignificant contribution of MERS-CoV (R^2^ = 0.02) and MERS (R^2^ = 0.06) to learning COVID-19 despite being a trending term in the early period of the SARS-CoV-2 outbreak and was initially surfacing in WHO’s pronouncements (Figure 4; WHO, 2020). On the use trend, as WHO changed the name of the novel coronavirus (nCoV) from 2019-nCoV to SARS-CoV-2, and naming the disease, COVID-19, the use of the previous title as search term also nosedived, thus indicating the reflection of Google Trends data to the reality, thus unveiling the trending events.

#### 5.1.2 What Terms Fueled Misinformation and Conspiracy Theories about COVID-19?

The literature identifies eight (8) terms representing the keywords denoting misinformation and conspiracy theories (Table 1). The misinformation are as follows: COVID-19 is just like “Common Cold (cold2020;” “ingesting or injecting bleach” can cure COVID-19 infection/kill the virus; COVID-19 is a hoax. Similarly, the conspiracy theories include, “5G” technology contributes to spreading COVID-19; COVID-19 is “biological weapon;” COVID-19 is “China Virus that was intentionally created;” and “COVID-19 was deliberately released.” As people learned about the science, concept, and structure of COVID-19, using the keywords that explains the coronavirus and the disease, they also learned the misinformation and conspiracy theories about the pandemic.

The scatter plot and trends analyses based on these search terms show that only two of the eight misinformation and conspiracy theories attracted significant attention of learners, namely, “common cold (Cold2020)” and “ChinaVirus.” In contrast, the other six (6) terms did not yield significant interest based on the worldwide Google Trends search index data. The coefficient of determination (R^2^) for the variables with R^2^ >= 10 include, Common Cold (cold2020): R^2^ =0.10; ChinaVirus: R^2^ =0.15. (Figure 5) shows the R^2^ for all the variables. The conspiracy theory that the virus was deliberately released did not have sufficient data on Google trends.

**Figure 5.**
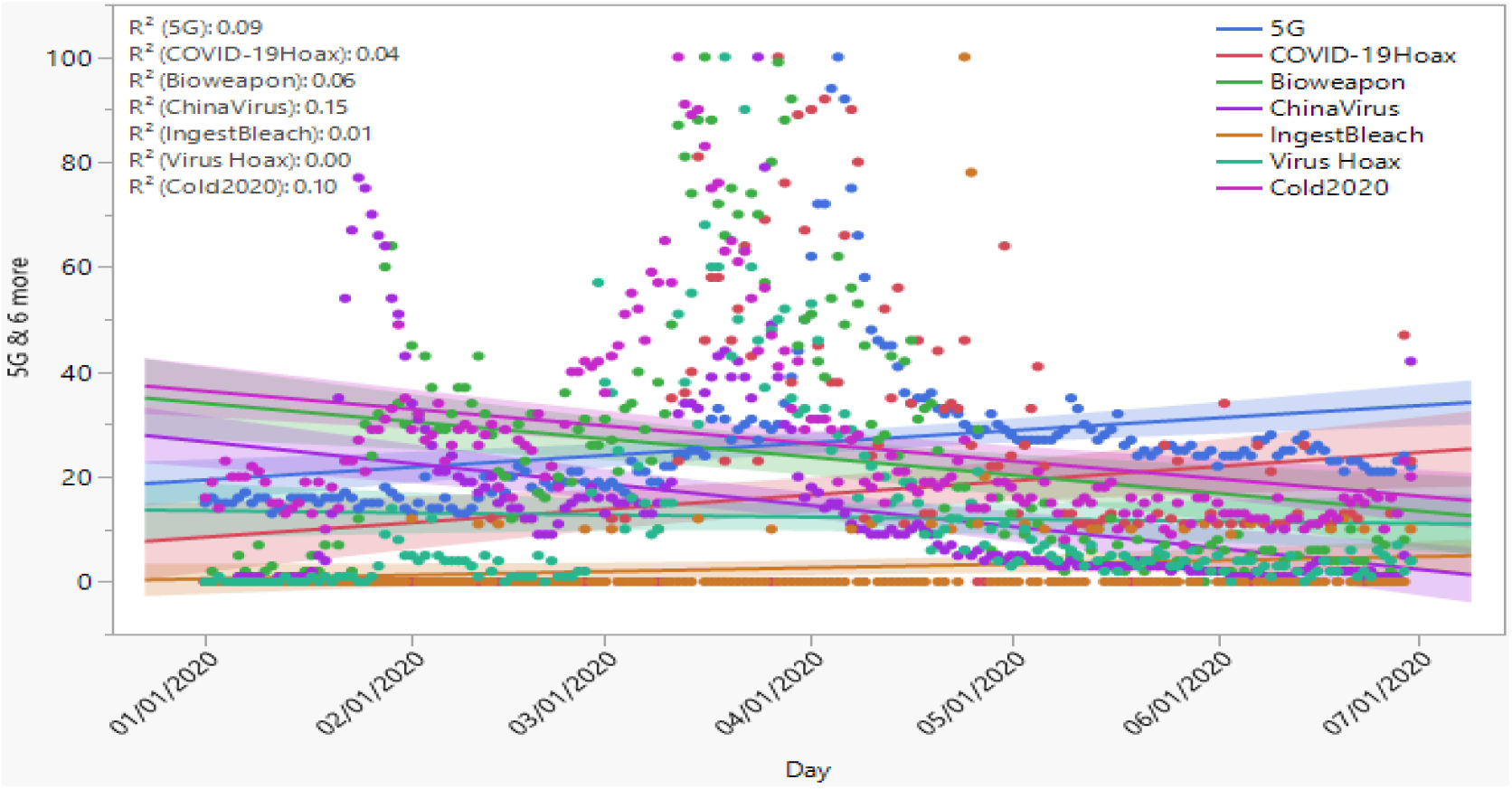
Monthly worldwide search index showing the learning terms that represent misinformation and conspiracy theories about COVID-19.

Based on the results shown, we selected the two search terms, the “Common Cold (cold2020)” as the misinformation and the “ChinaVirus” conspiracy theory for further analysis on how the two variables impact the learners’ behavior towards the public health guidelines (Section 5.2).

#### 5.1.3 Public Health Safety Measures

As discussed in Section 2.3, COVID-19 has lasted several months, but there is no specific cure yet. However, experts emphasize the need for everyone to practice and abide by the recommended public health safety measures of social distancing, wearing a facial mask and handwashing as ways to limit the human-to-human spread of COVID-19. Similarly, those exposed to imminent risks of COVID-19 infection are required to isolate or quarantine (Matias, Dominski, & Marks, 2020; Miller et al., 2020a; 2020b). This section investigates the influence of what people learned about SARS-CoV-2 and COVID-19 through a web search on their behaviors towards public health safety measures. Figure 6 shows the search index trends.

**Figure 6:**
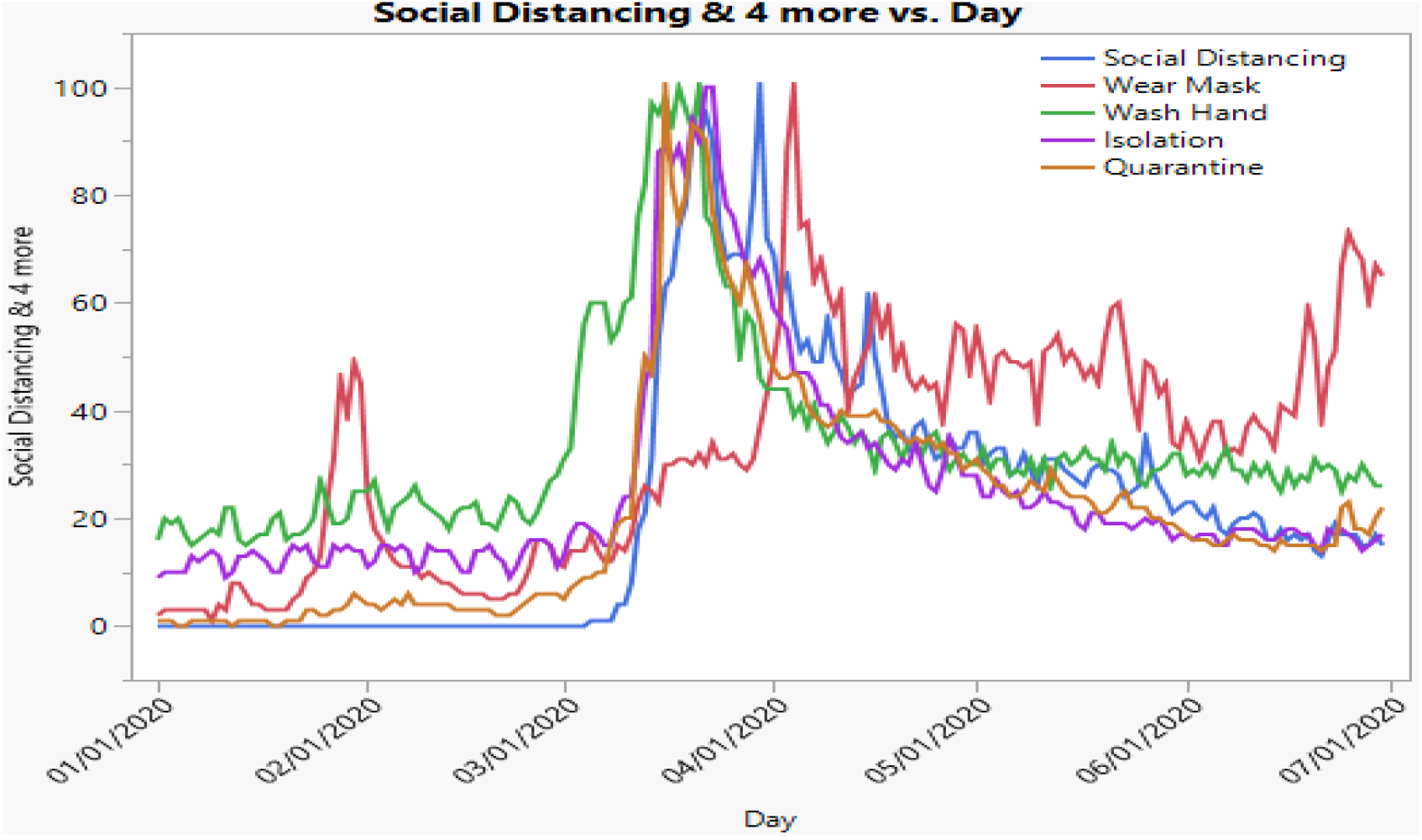
Daily Search Index to learn about the public health safety measures.

The trend analysis of the daily web search index of the terms that people used to learn about public health safety measures (Figure 7) highlights two things. First, the results show that people did not start learning about the public health measures in the early days of the COVID-19 outbreak until 3-4 months, occurring in the first wave of the COVID-19 infection with increasing fatality. It does appear that people only began learning about the pandemic as COVID-19 infections skyrocket, indicating a late response. This trend is consistent with the literature, which attributes the late response to the lag in public health communication, which contributed to the spread of misinformation and conspiracy theories, with significant negative impacts on the peoples’ reactions to the public health measures (Taylor, 2020). Second, only two (2) out of the five (5) public health safety measures attracted the interest of learners through a web search, namely, wearing a facial mask (R^2^ =0.56) and, to a lesser extent, social distancing (R^2^=0.13). The coefficient of determination and the correlation shows that only a few people bothered to learn other public health safety measures (handwashing, isolation, and quarantine), all having R^2^ ranging from 0.01 to 0.09.

**Figure 7:**
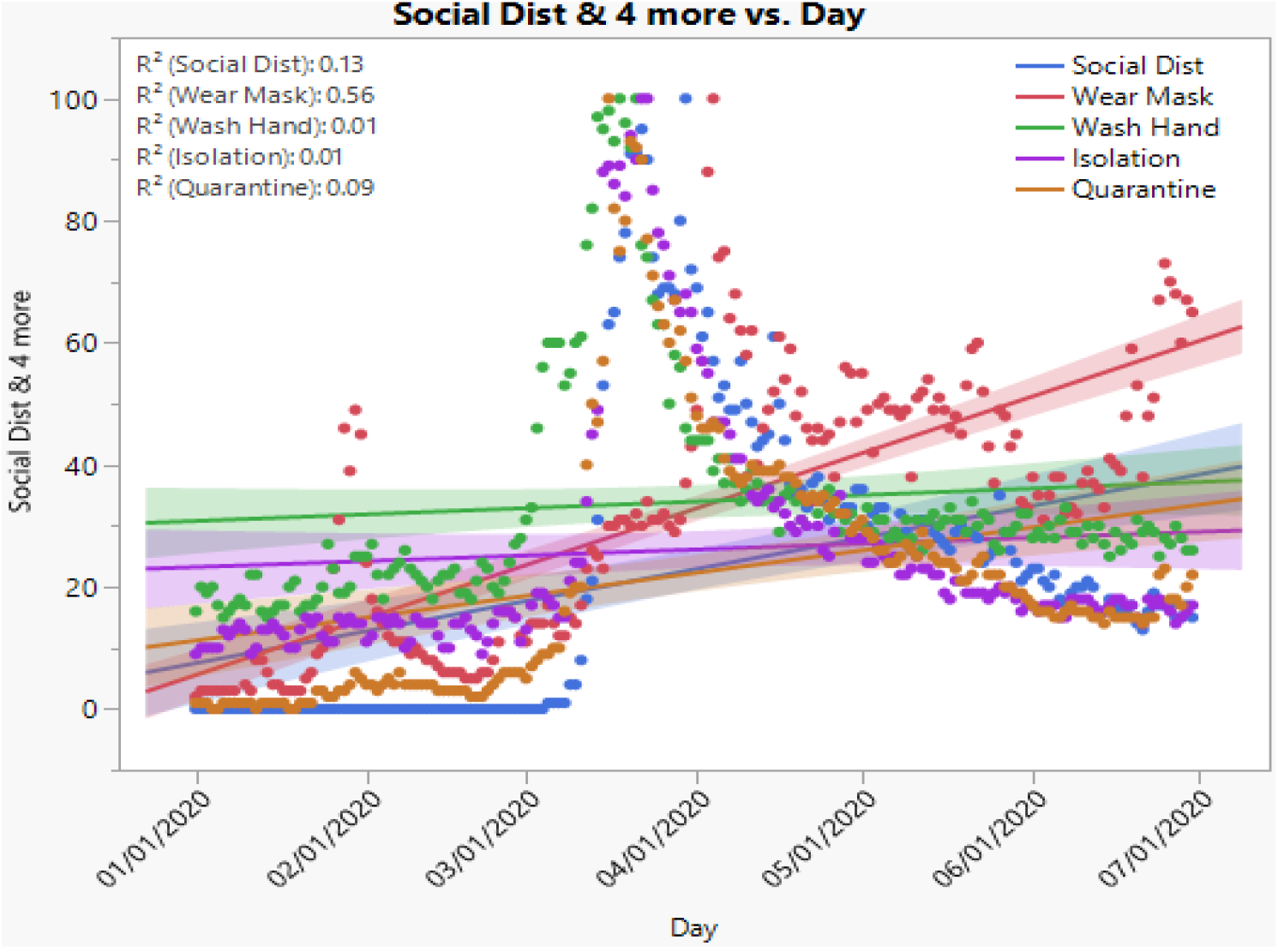
The trend analysis showing the public’s interest in learing about public health safety measures 5.2 Hypotheses Testing

### 5.2 Hypotheses Testing

The results presented in Section 5.1 show the web search index of twenty-three (23) variables or search terms that people used to learn or gain information about SARS-CoV-2 and COVID-19, the misinformation and conspiracy theories, and public health safety measures. However, some of the variables did not contribute meaningfully to peoples’ learning or gained information about the pandemic and were removed from further analysis (Figures 4, 5, 7). This section presents further analysis involving the selected variables that contributed to learning about SARS-CoV-2 and COVID-19 based on the determination coefficient (R^2^ >= 10). Six (6) of the variables relate to learning about the concept, structure, and science of coronavirus pandemic, two (2) relate to misinformation and conspiracy theory, and the last two learning terms address the public health measures (wearing of facial mask and social distancing). Figure 8 shows the list of variables selected for further analyses.

**Figure 8.**
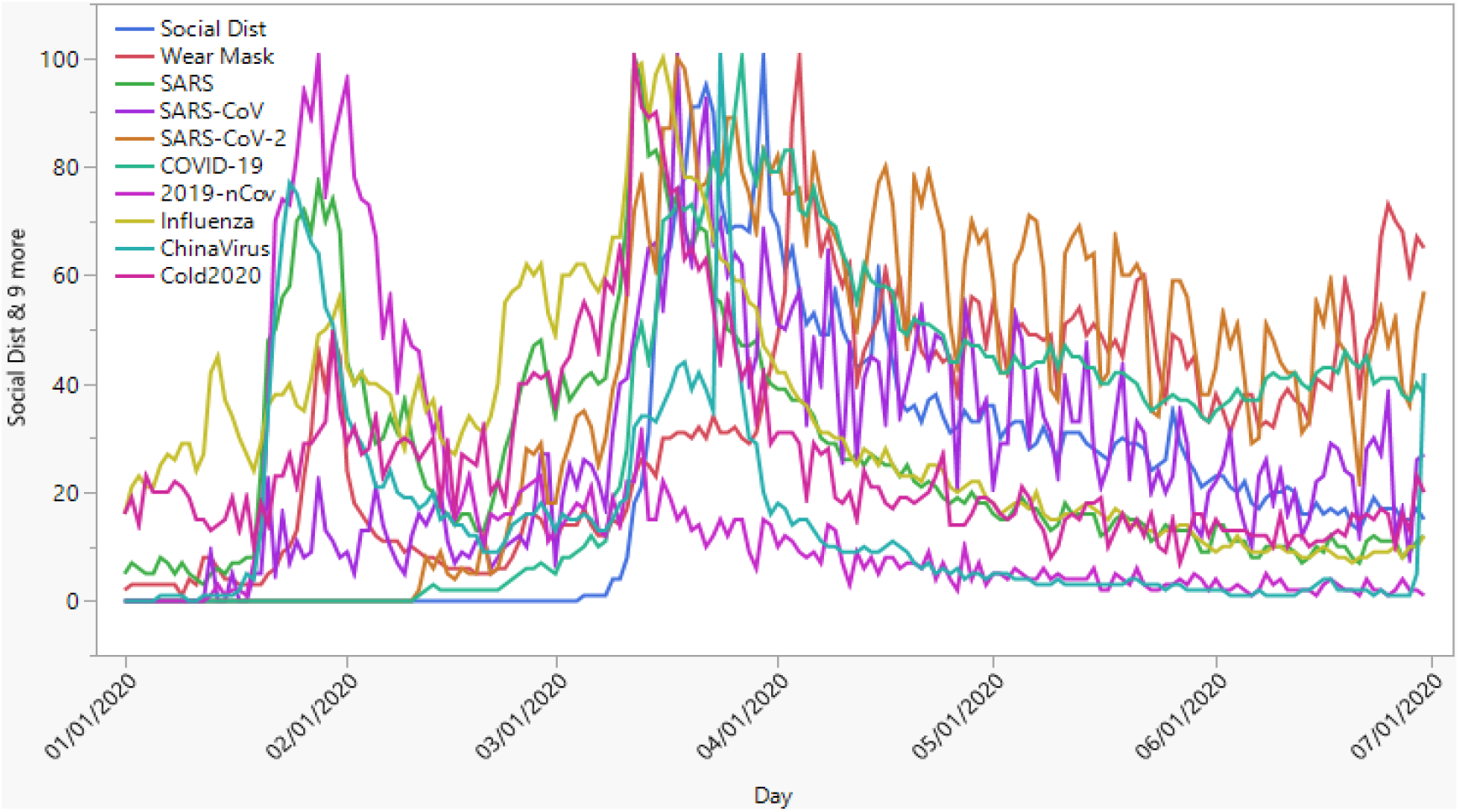
The usage trends of ten variables that contributed to learning about SARS-CoV-2 and COVID-19

The results show three essential highlights from the daily use of the ten learning terms. First, the most popular search terms used at the initial outbreak of the pandemic in early January 2020 were “common cold” and “China Virus,” representing the misinformation and conspiracy theory, and Influenza, a previous pandemic (Table 1). Other popular learning terms include SARS-CoV (a previous coronavirus outbreak) and 2019-nCoV (the initial name of SARS-CoV-2). The other learning terms (SARS-CoV-2 and COVID-19) were not used until February 2020, when WHO renamed the coronavirus from 2019-nCoV to SARS-CoV-2. Figure 8 shows further details about the time and pattern of use of the variables.

#### 5.2.1 What Did People Learn about SARS-CoV-2 and COVID-19 Through Web Search?

Figures 8 and 9 present the results of what people learned about the ongoing COVID-19 pandemic. The search keywords are approximations to what people learned or the knowledge gained about COVID-19.

**Figure 9.**
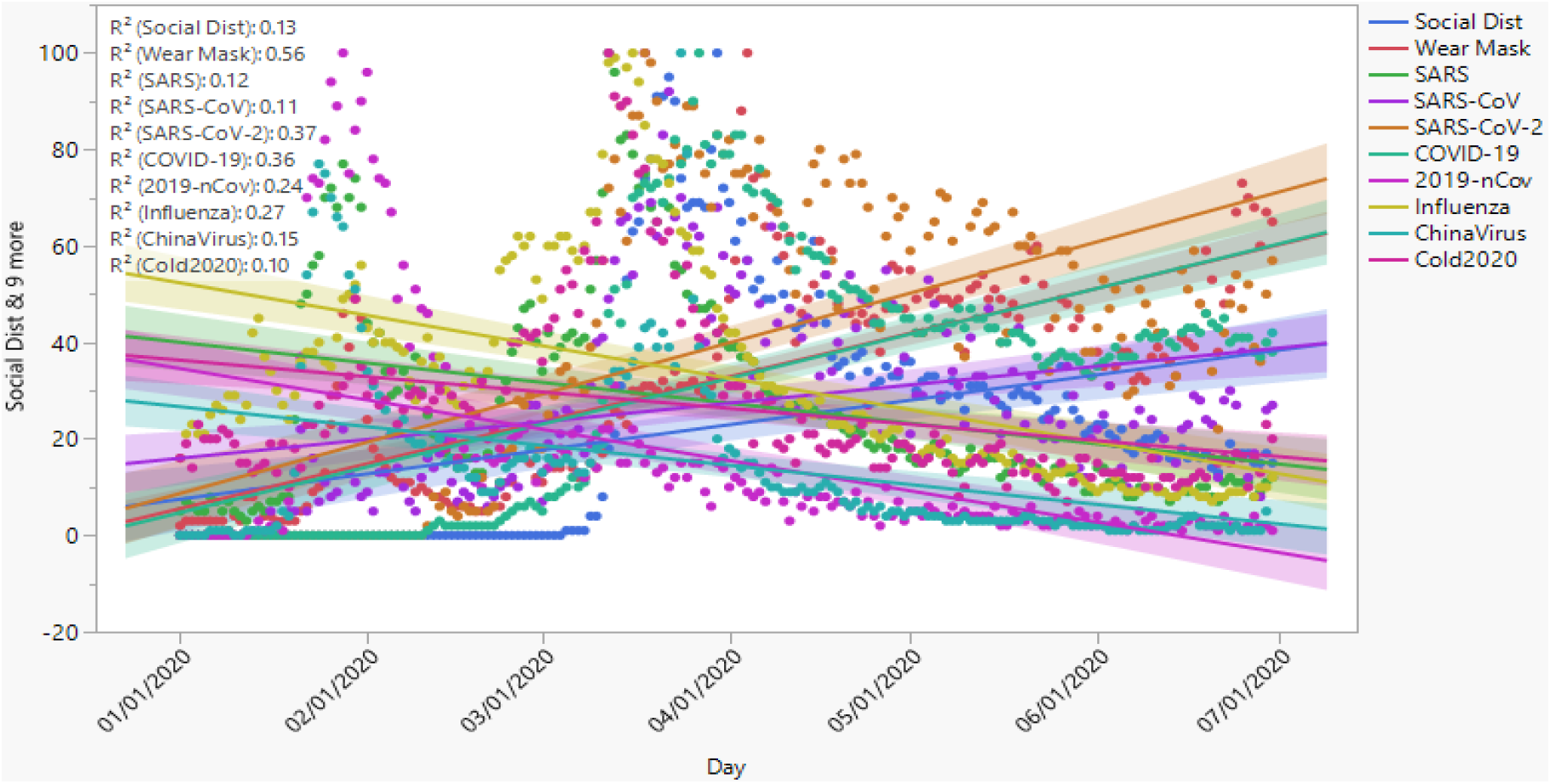
The variables and contributions to learning about SARS-CoV-2 and COVID-19 (6), misinformation and conspiracy theories (2), and public health measures (2)

The coefficient of determination (R2) shows that the most effective learning term or phrase is “wearing a facial mask” (R^2^ =0.56), the expression that explains one of the public health measures to limit the spread of COVID-19. The result indicates that, of the people who used learned information about the ongoing pandemic, a significant number obtained information about the public health safety guidelines on wearing a facial mask. Simultaneously, many people also got information about the concept, structure, and science of SARS-CoV-2 (R^2^ =0.37), 2019-nCoV (R^2^ =0.24), and the coronavirus disease, COVID-19 (R =0.36). Other related subjects that people learned while obtaining information about SARS-CoV-2 and COVID-19 were SARS (R^2^ = 0.12) and SARS-CoV (R^2^ = 0.11), which relate to past epidemics but are closely related to the ongoing pandemic.

However, the association among the variables was more bullish. For example, the correlation analysis shows a robust, positive, and significant direct association between SARS-CoV and SARS-CoV-2 (r=0.91), SARS-CoV, and COVID-19 (r=0.88), and SARS and 2019-nCoV (r=0.85). Table 2 shows a complete result and Figure 10 highlights visually the significant associations among variables. The overall results indicate that a good number of people indeed learned about SARS-CoV-2 and COVID-19. Based on these results, we can reject the null hypothesis (H1_0_) and accept the alternative view, concluding that web search using the correct terms helped people gain information about the ongoing COVID-19 pandemic.

**Table 2.** Correlation matrix showing the relationships among the learning terms.

|  | 1 | 2 | 3 | 4 | 5 | 6 | 7 | 8 | 9 | 10 |
| --- | --- | --- | --- | --- | --- | --- | --- | --- | --- | --- |
| 1. SARS-CoV-2 | 1.00 |  |  |  |  |  |  |  |  |  |
| 2. COVID-19 | 0.95 | 1.00 |  |  |  |  |  |  |  |  |
| 3. 2019-nCoV | -0.045** | -0.073** | 1.00 |  |  |  |  |  |  |  |
| 4. SARS-CoV | 0.91 | 0.88 | 0.13** | 1.00 |  |  |  |  |  |  |
| 5. SARS | 0.28 | 0.25 | 0.85 | 0.45 | 1.00 |  |  |  |  |  |
| 6. Influenza | -0.01** | -0.04** | 0.73 | -0.14** | 0.79 | 1.00 |  |  |  |  |
| 7. Cold2020<br>(Common Cold) | 0.10** | 0.06** | 0.69 | 0.23 | 0.78 | 0.86 | 1.00 |  |  |  |
| 8. ChinaVirus | 0.13** | 0.11** | 0.88 | 0.30 | 0.92 | 0.78 | 0.77 | 1.00 |  |  |
| 9. Social<br>Distancing | 0.93 | 0.94 | -0.12** | 0.87 | 0.23* | -0.06** | 0.01** | -0.08** | 1.00 |  |
| 10. Wear Mask | 0.66 | 0.68 | -0.21* | 0.61 | -0.01** | -0.43 | -0.27 | -0.12** | 0.67 | 1.00 |

\* $p \leq 0.05$ ; \*\*Non-significant association with $p > 0.05$ ; All other associated variables are significant at $p < 0.001$

**Figure 10.**
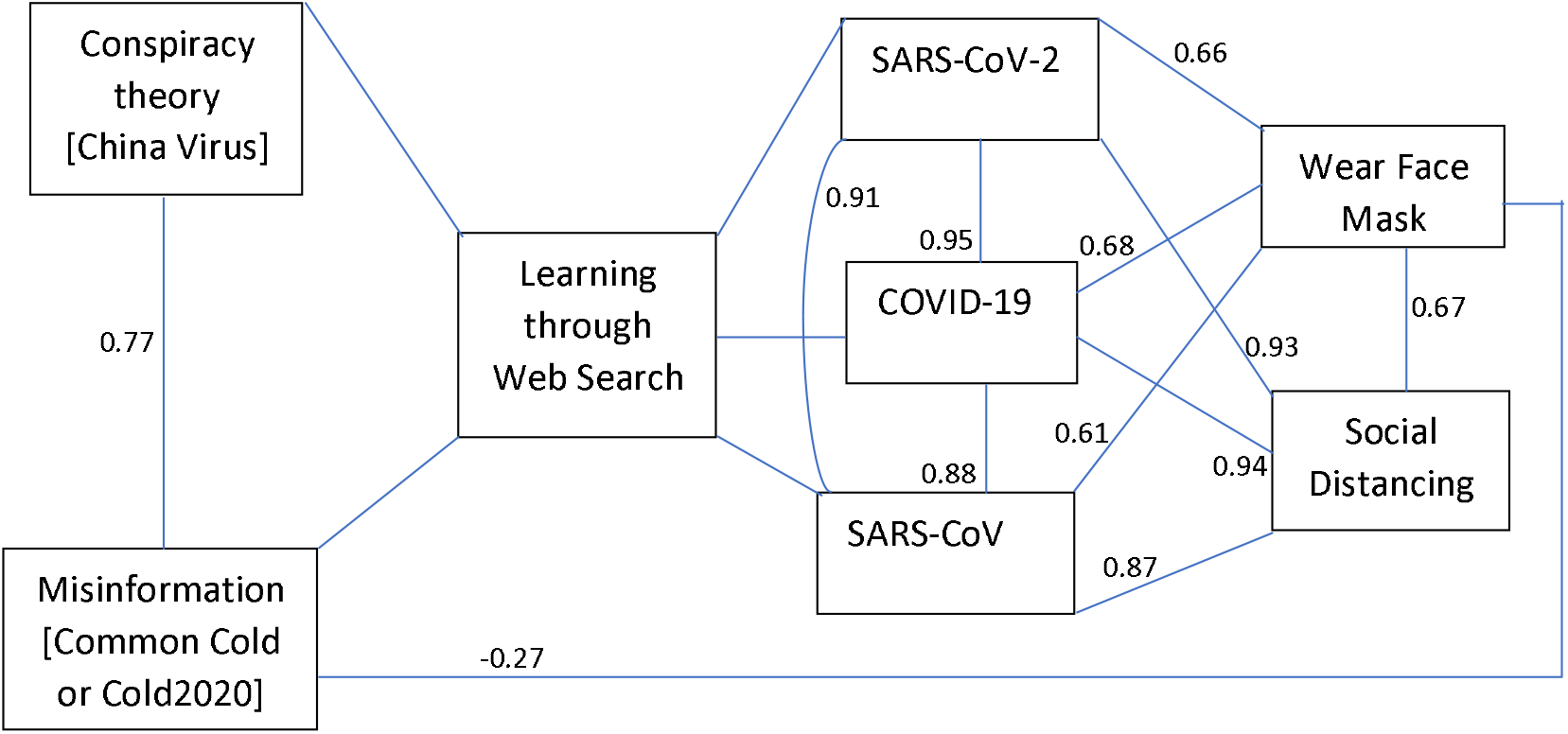
Visual representation of the significant associations among the variables (p<0.001).

Another highlight of the study relates to a previous pandemic named Influenza H1N1 (R^2^ = 0.27). As the concept, structure, and science of Influenza is entirely different from SARS-CoV-2, using the term does not help users to learn about the ongoing pandemic.

The results (Table 2 above and the visualized correlation matrix, Figure 10) show that despite the high search index (average = 32.3 points, even higher than the search index for COVID-19), using the term does not lead to learning about the ongoing pandemic based on the coefficient (r=0.01), which was also not significant at p=0.05. Influenza as a learning term does not enhance learning about COVID-19.

#### 5.2.2 Information Learned Versus Behavior Towards Public Health Behaviors

This section examines the relationship between what people learned and attitude toward public health measures. The results of the correlation analysis show a strong positive association between SARS-CoV-2 and “wearing a facial mask” (r=0.66) and SARS-CoV-2 and social distancing (r=0.93). Similarly, the learning term (COVID-19) has a strong positive relationship with wearing a facial mask (r=0.94) and COVID-19 and social distancing (r=0.68). The previous coronavirus epidemic (SARS-CoV) also has a strong positive relationship with wearing facial mask and social distancing (r=0.61 and 0.87, respectively). Thus, the more people learned about the SARS-CoV-2 and COVID-19, the more they pay attention to the recommended public health measures of wearing a facial mask and social distancing. Conversely, learning about Influenza, misinformation, and conspiracy theories results in less interest in learning the public health safety guidelines towards wearing a facial mask and social distancing. Based on these results, we test the hypothesis (H2), rejecting the null hypothesis and accept the alternative view that there is an association between what people learned about COVID-19 and their behavior towards the recommended public health measures Table 2, Figure 10).

#### 5.2.3 Misinformation, Conspiracy Theory, and Public Health Measures

This section examines the relationship between misinformation, conspiracy theories, and public health measures using the correlation analysis between what people learned through a web search. As discussed earlier, only two of the five search terms representing misinformation and conspiracy theory met the threshold of R^2^ >= 10 and got selected for further analysis. We use the results to test hypotheses H3A and H3B. Hypothesis H3A focuses on the effect of misconception and public health guidelines, while H3B addresses the impact of conspiracy theory on people’s behavior towards public health safety guidelines. The results of the correlation analysis show that there is a strong and significant inverse relationship between misinformation (Common Cold labeled cold2020) and public health measure of wearing a facial mask (r=-0.60; p<0.001). Also, there is an inverse association between misinformation (Common Cold) and public health guideline of social distancing (r=-0.27; p<0.001). On this note, we reject the null hypothesis and accept the alternative hypothesis that there is an association between misinformation and people’s response to the public health guidelines of wearing facial masks and social distancing as measures to limit the spread of COVID-19.

On the association between the conspiracy theory (ChinaVirus) and public health measure of wearing a facial mask, there is also an inverse association based on the correlation coefficient (r=-0.12; p>0.05). A similar inverse relationship between the conspiracy theory (ChinaVirus) and the public health measure of social distancing (r=-0.08; p>0.05). However, the associations were not statistically significant in the case of the conspiracy theory (ChinaVirus). In this regard, we accept the null hypothesis that there is no association between conspiracy theory and people’s response to the public health guidelines of wearing facial masks and social distancing as measures to limit the spread of COVID-19.

## 6. Conclusion

This study offers empirical results showing that a good portion of the global population learned about the outbreak of SARS-CoV-2 and COVID-19 through a web search particularly in the early period of the pandemic. The period covers the initial days, weeks, and months from the emergence of the novel coronavirus in December 2019 up to June 30, 2019, when the public became more aware of the pandemic, especially after the first wave (Taylor, 2020).

Further, people used the web to learn about the ongoing COVID-19 in three aspects, knowledge about the novel coronavirus (2019-nCoV, later renamed SARS-CoV-2), misinformation and conspiracy theories, and obtaining information about the public health safety measures. The more people focused attention on learning about SARS-CoV-2 and COVID-19, the more they also learned about the public health measures, and vice versa. On the other hand, those who focused attention on learning about misinformation and conspiracy theories had little or no interest in finding information about wearing a facial mask or social distancing. Although an aspect of two studies (Sulyok, Ferenci, & Walker, 2020; Rovetta, & Bhagavathula, 2020) examined the use of the web to search about COVID-19, the limit the study to selected provinces in Italy and how the search impacts the infection rates of COVID-19 in those areas. Therefore, this study is the first to examine not only what people learned but also how these influence people’s social behavior towards the public health safety guidelines.

The conclusions from this study corroborate the real-life behaviors of people about the ongoing COVID-19 pandemic. For example, people who have a proper understanding of the concept, structure, and science of COVID-19 are more likely to comply with public health safety measures. On the other hand, those who echo the misinformation or conspiracy theories are more likely to exhibit defiant behaviors against the public health safety measures (Miller et al., 2020a; 2020b; Allington et al., 2020).

## Data Availability

Not applicable

## Notes

### Competing Interest Statement

The authors have declared no competing interest.

